# Acute Myocardial Injury of Patients with Coronavirus Disease 2019

**DOI:** 10.1101/2020.03.05.20031591

**Authors:** Huayan Xu, Keke Hou, Hong Xu, Zhenlin Li, Huizhu Chen, Na Zhang, Rong Xu, Hang Fu, Ran Sun, Lingyi Wen, Linjun Xie, Hui Liu, Kun Zhang, Joseph B. Selvanayagam, Chuan Fu, Shihua Zhao, Zhigang Yang, Ming Yang, Yingkun Guo

**Affiliations:** Department of Radiology, Key Laboratory of Birth Defects and Related Diseases of Women and Children of Ministry of Education, West China Second University Hospital, Sichuan University, Chengdu, China; Department of Radiology, Public Health Clinical Center of Chengdu, Cheng Du, China; Department of Radiology, State Key Laboratory of Biotherapy, West China Hospital, Sichuan University, Chengdu, China; Department of Radiology, Cardiac Imaging Center, Fuwai Hospital, National Center for Cardiovascular Diseases of China, Chinese Academy of Medical Sciences and Peking Union Medical College, Beijing, China; Department of Respiratory Medicine, Public Health Clinical Center of Chengdu, Cheng Du, China; Department of Cardiovascular Medicine, Flinders Medical Centre, Flinders University of South Australia, Adelaide, Australia; Department of Ultrasonography, West China Second University Hospital, Sichuan University, Chengdu, China

## Abstract

**Background:** Since the outbreak of the Coronavirus Disease 2019 (COVID-19) in China, respiratory manifestations of the disease have been observed. However, as a fatal comorbidity, acute myocardial injury (AMI) in COVID-19 patients has not been previously investigated in detail. We investigated the clinical characteristics of COVID-19 patients with AMI and determined the risk factors for AMI in them.

**Methods:** We analyzed data from 53 consecutive laboratory-confirmed and hospitalized COVID-19 patients (28 men, 25 women; age, 19–81 years). We collected information on epidemiological and demographic characteristics, clinical features, routine laboratory tests (including cardiac injury biomarkers), echocardiography, electrocardiography, imaging findings, management methods, and clinical outcomes.

**Results:** Cardiac complications were found in 42 of the 53 (79.25%) patients: tachycardia (n=15), electrocardiography abnormities (n=11), diastolic dysfunction (n=20), elevated myocardial enzymes (n=30), and AMI (n=6). All the six AMI patients were aged >60 years; five of them had two or more underlying comorbidities (hypertension, diabetes, cardiovascular diseases, and chronic obstructive pulmonary disease). Novel coronavirus pneumonia (NCP) severity was higher in the AMI patients than in patients with non-definite AMI (p<0.001). All the AMI patients required care in intensive care unit; of them, three died, two remain hospitalized. Multivariate analyses showed that C-reactive protein (CRP) levels, NCP severity, and underlying comorbidities were the risk factors for cardiac abnormalities in COVID-19 patients.

**Conclusions:** Cardiac complications are common in COVID-19 patients. Elevated CRP levels, underlying comorbidities, and NCP severity are the main risk factors for cardiac complications in COVID-19 patients.

## Introduction

The widespread outbreak of the Coronavirus Disease 2019 (COVID-19, previously known as 2019-nCoV) in China is a big challenge for public health and medical care. Since its first emergence in Wuhan city in late December 2019, COVID-19 has already spread to 26 countries. As of March 02, 2020, there were over 87137 confirmed cases worldwide, with 2977 deaths.^1^ To date, several latest studies have reported the clinical characteristics of hospitalized patients with 2019 novel coronavirus pneumonia (NCP), including signs, symptoms, laboratory test results, imaging features, therapeutic strategies and effects, and multiple organ dysfunction.^2-5^ Notably, acute myocardial injury (AMI), defined as troponin T-hypersensitivity (TNT-HSST) serum levels > 99th percentile upper reference limit (>28 pg/ml) by the American College of Cardiology/American Heart Association Task Force for myocardial infarction and non-myocardial infarction diseases,^6^ was detected in approximately 7.2%–12% COVID-19 patients in previous studies.^2-3^ Furthermore, both severe acute respiratory syndrome (SARS) and Middle East respiratory syndrome (MERS) have been linked to acute myocarditis, AMI, and rapid-onset heart failure.^7-9^ Reportedly,^2^ nearly 40% of hospitalized patients with confirmed COVID-19 have underlying comorbidities such as cardiovascular or cerebrovascular diseases. Furthermore, among COVID-19 patients, those with underlying cardiovascular diseases can be more severely affected and may have more adverse outcomes than those without underlying diseases. Thus, undoubtedly, special medical attention should be paid to COVID-19 patients with cardiac complications and underlying cardiovascular diseases. On February 13, 2020, the ACC released an ACC Clinical Bulletin titled “Cardiac Implications of Novel Wuhan Coronavirus (COVID-19)”^10^ for addressing cardiac implications of this disease and offering early clinical guidance given the current uncertainty over COVID-19. However, detailed clinical characteristics of COVID-19 patients with cardiac involvement, especially those with AMI, have not been investigated in detail. Thus, we aimed to describe the clinical characteristics of COVID-19 patients with AMI and to further determine the risk factors for AMI in them.

## Methods

### Study population

The institutional ethics board of our institutes approved this study (No. 2020.43). We retrospectively enrolled 53 laboratory-confirmed COVID-19 patients hospitalized between January 02, 2020 and February 14, 2020. The patients were diagnosed and classified according to the World Health Organization interim guidance.^7^ NCP types were classified as mild, common, severe, and critically severe according to the COVID-19 patient management guideline issued on February 8, 2020.^11^

### Procedures

Data on epidemiological and demographic characteristics, clinical signs and symptoms, underlying comorbidities, complications, and treatment management (such as antiviral or antibiotic therapy, respiratory support, continuous renal replacement therapy, and extracorporeal membrane oxygenation [ECMO]) were collected from the patients’ medical records. Laboratory test results including those of blood routine tests, C-reactive protein (CRP) and D-dimer tests, and blood gas analysis were recorded and compared. The levels of cardiac injury markers including lactate dehydrogenase (LDH), hydroxybutyrate dehydrogenase, creatine kinase (CK), CK-MB, myoglobin (MYO), amino N-terminal pro-brain natriuretic peptide, and TNT-HSST were determined at admission. The AMI group was defined as patients with TNT-HSST serum levels > 99th percentile upper reference limit (>28 pg/ml).^6^ The group with cardiac marker abnormalities but non-definite AMI was defined as patients with TNT-HSST serum levels < 28pg/ml but increase in the levels of any of the other abovementioned cardiac markers. The group without cardiac marker abnormalities comprised patients without elevation in the levels of any of the cardiac markers. Chest computed tomography (CT) was performed on the day of admission. The period of illness onset to admission was defined from the day when signs or symptoms were first noticed to the day of admission. All enrolled patients underwent two-dimensional echocardiography (2D echo) and electrocardiography (ECG) during hospitalization. Acute respiratory distress syndrome (ARDS) was identified according to the Berlin definition^12^ and acute kidney injury (AKI) according to the Kidney Disease Improving Global Outcomes definition.^13^

### Statistical analysis

Statistical analysis was performed using SPSS version 21.0 (Armonk, NY; Graphpad version 7.00, San Diego, CA). Data were presented as median (interquartile range; IQR) if non-normally distributed and as mean (standard deviation; SD) if normally distributed. Categorical variables were presented as n (%). Kruskal–Wallis analysis, Chi-square test, and one-way analysis of variance were used for multiple group comparisons according to data characteristics. Bivariate correlation was calculated using Pearson or Spearman method as appropriate to detect the potential risk factors for AMI. In all analyses, two-tailed p-values of <0.05 were considered significant.

## Results

### Baseline data

The baseline data of the 53 COVID-19 patients are listed in **Table 1**. Of them, six (11.32%) patients had AMI (all aged >60 years; median age, 78.5 years [IQR 60.5, 81.75]); 24 (45.38%) patients were defined as the cardiac marker abnormalities group (6 [25%] aged >60 years; median age, 48.50 years old [IQR 37.00, 62.75]). Cardiac markers of 23 (43.39%) patients were within the normal range (4 [17.39%] aged >60 years; median age, 40.00 years [IQR 23.00, 51.00]). There were no significant gender differences among the three subgroups. Epidemiology of all the groups was similar (all p>0.05), such as being a local resident, Wuhan residence exposure, and COVID-19 exposure. No patients had direct Huanan seafood market exposure. No exact epidemiological link was found between the first case and the later ones. Almost all the patients’ first onset symptoms were fever (n=47, 88.68%) and dry cough (n=41, 77.36%). Furthermore, the mean systolic blood pressure of the AMI patients was higher than those of the other two groups (p<0.001). Fifteen (28%) of all the patients had tachycardia.

**Table 1.**
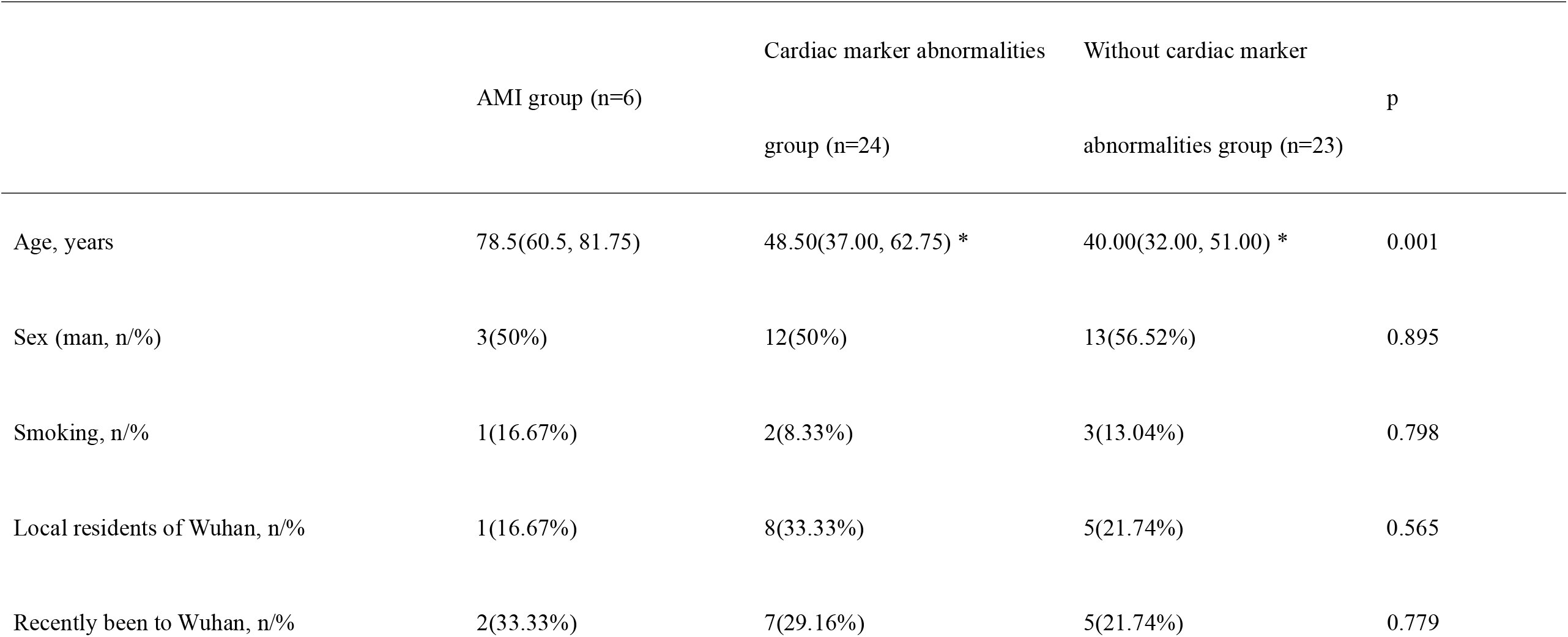
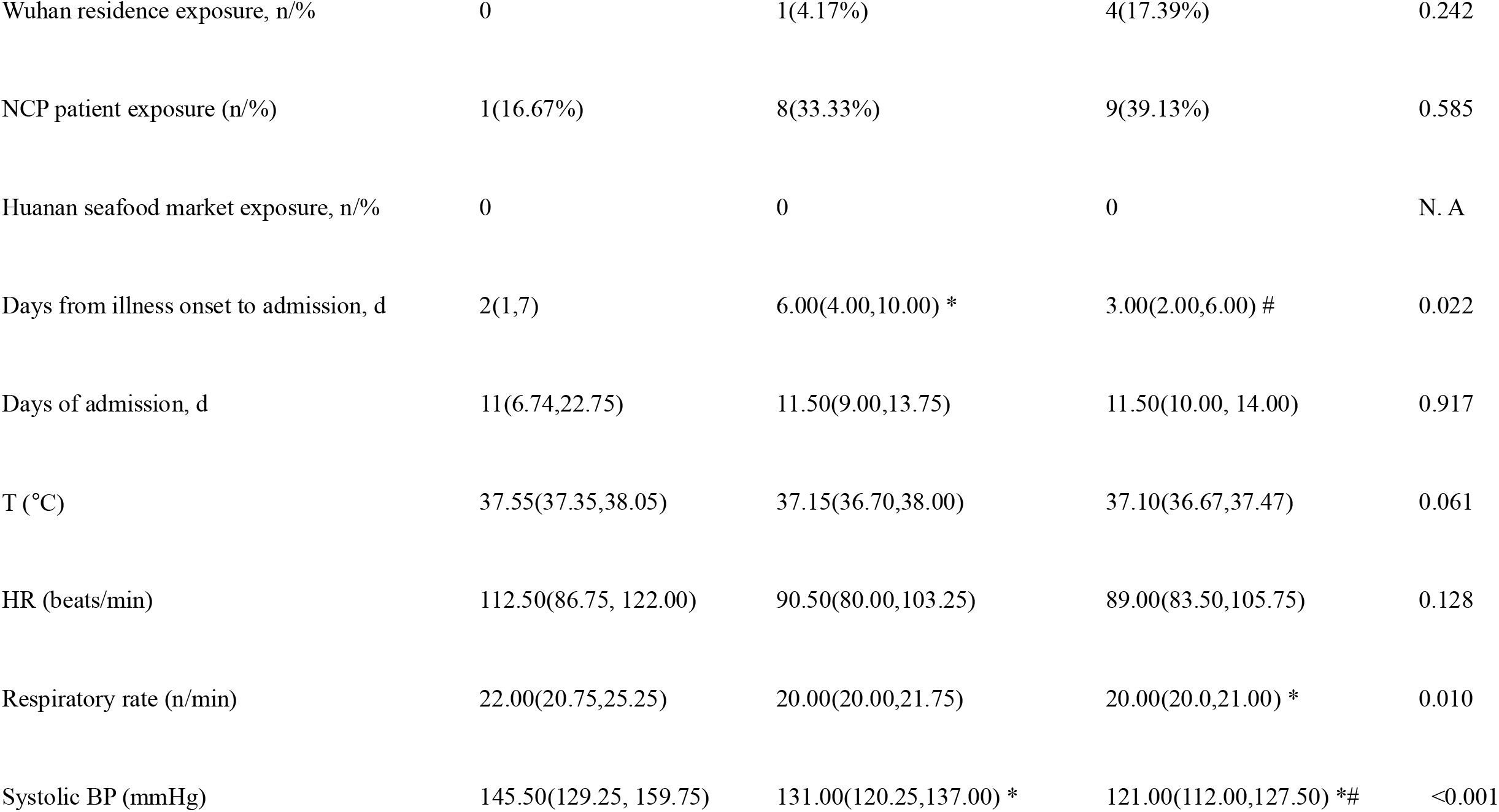
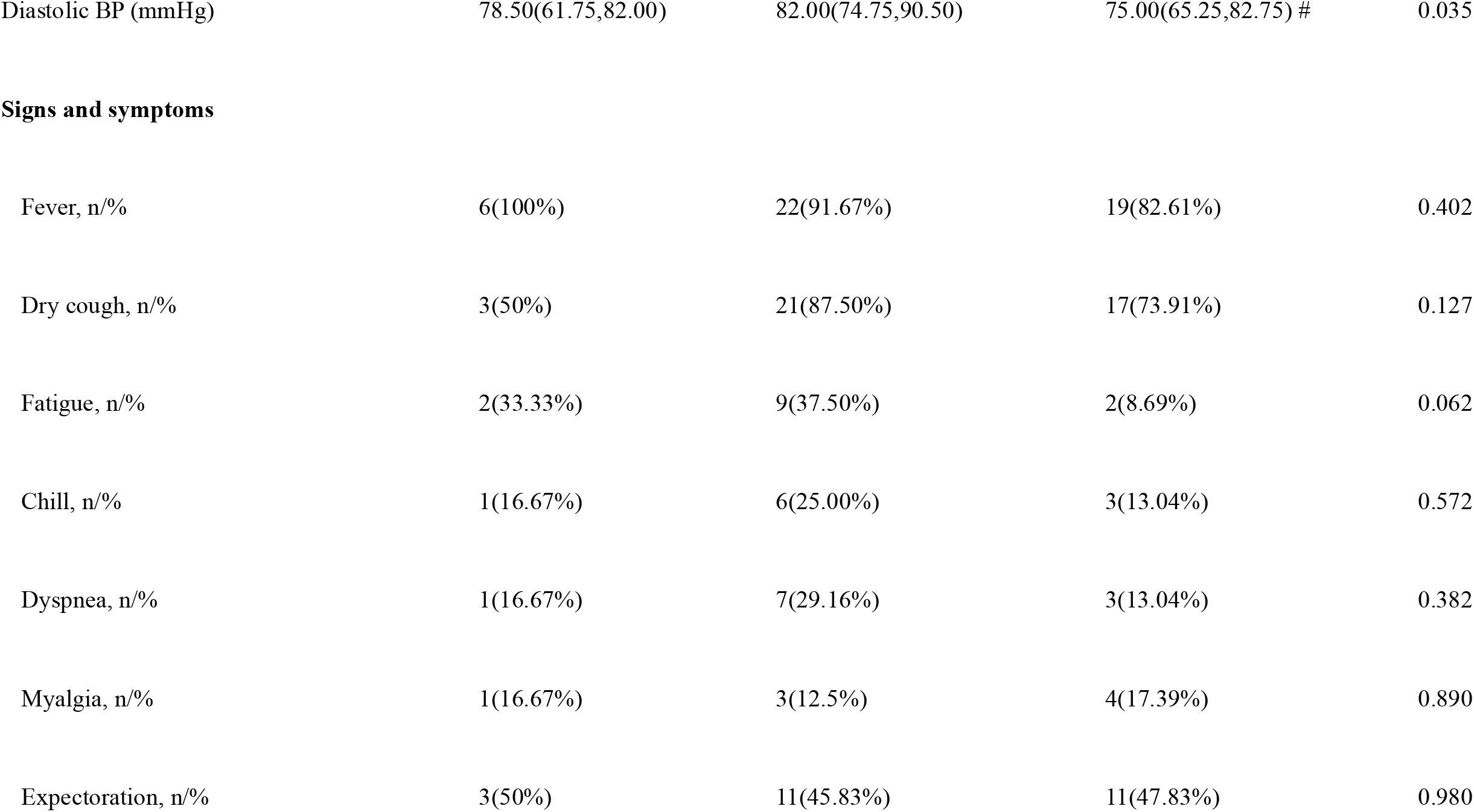
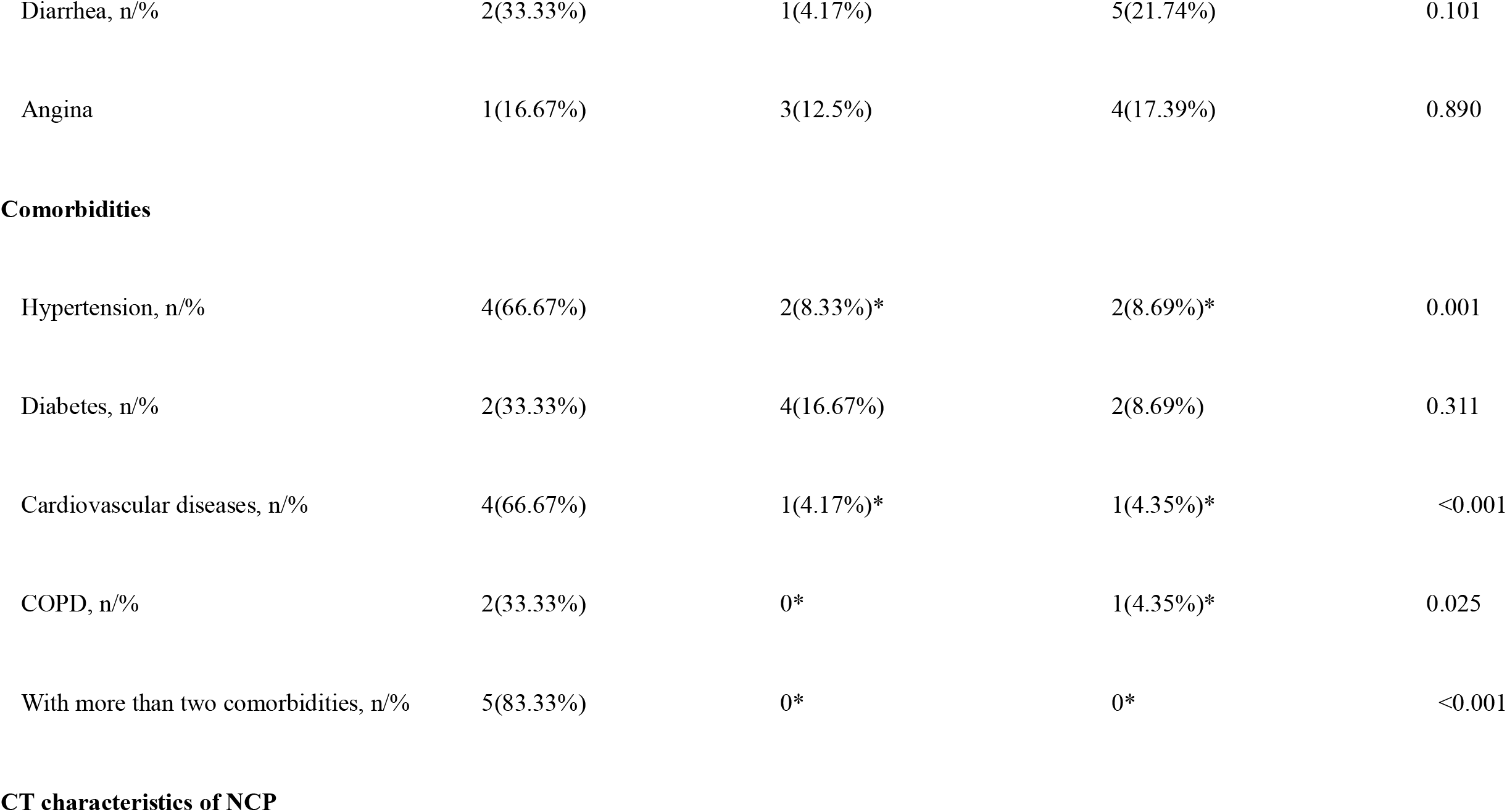
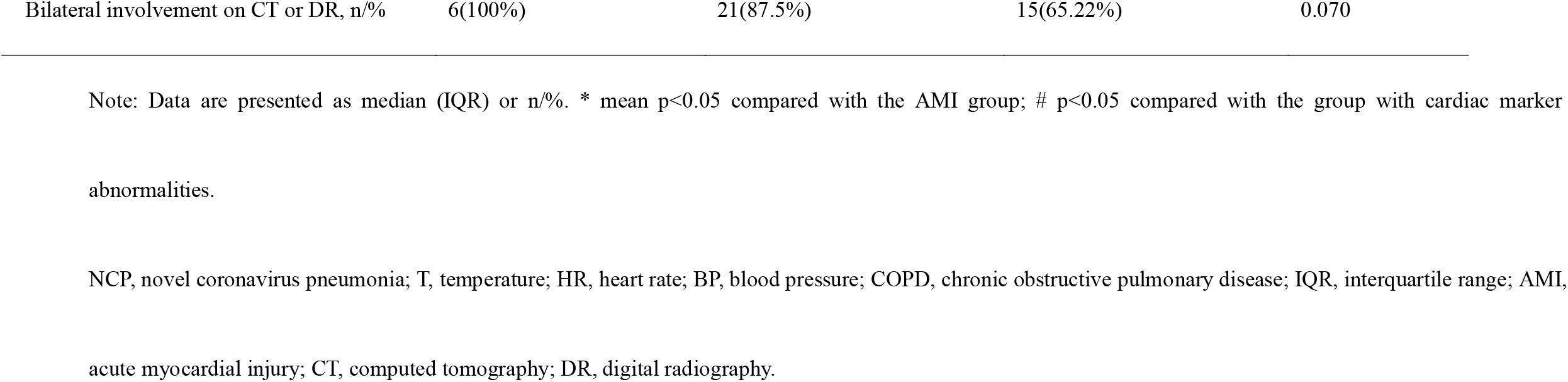
Baseline characteristics.

|  | AMI group (n=6) | Cardiac marker abnormalities<br>group (n=24) | Without cardiac marker<br>abnormalities group (n=23) | p |
| --- | --- | --- | --- | --- |
| Age, years | 78.5(60.5, 81.75) | 48.50(37.00, 62.75) * | 40.00(32.00, 51.00) * | 0.001 |
| Sex (man, n/%) | 3(50%) | 12(50%) | 13(56.52%) | 0.895 |
| Smoking, n/% | 1(16.67%) | 2(8.33%) | 3(13.04%) | 0.798 |
| Local residents of Wuhan, n/% | 1(16.67%) | 8(33.33%) | 5(21.74%) | 0.565 |
| Recently been to Wuhan, n/% | 2(33.33%) | 7(29.16%) | 5(21.74%) | 0.779 |
| Wuhan residence exposure, n/% | 0 | 1(4.17%) | 4(17.39%) | 0.242 |
| NCP patient exposure (n/%) | 1(16.67%) | 8(33.33%) | 9(39.13%) | 0.585 |
| Huanan seafood market exposure, n/% | 0 | 0 | 0 | N. A |
| Days from illness onset to admission, d | 2(1,7) | 6.00(4.00,10.00) * | 3.00(2.00,6.00) # | 0.022 |
| Days of admission, d | 11(6.74,22.75) | 11.50(9.00,13.75) | 11.50(10.00, 14.00) | 0.917 |
| T (°C) | 37.55(37.35,38.05) | 37.15(36.70,38.00) | 37.10(36.67,37.47) | 0.061 |
| HR (beats/min) | 112.50(86.75, 122.00) | 90.50(80.00,103.25) | 89.00(83.50,105.75) | 0.128 |
| Respiratory rate (n/min) | 22.00(20.75,25.25) | 20.00(20.00,21.75) | 20.00(20.0,21.00) * | 0.010 |
| Systolic BP (mmHg) | 145.50(129.25, 159.75) | 131.00(120.25,137.00) * | 121.00(112.00,127.50) *# | <0.001 |
| Diastolic BP (mmHg) | 78.50(61.75,82.00) | 82.00(74.75,90.50) | 75.00(65.25,82.75) # | 0.035 |
| **Signs and symptoms** |  |  |  |  |
| Fever, n/% | 6(100%) | 22(91.67%) | 19(82.61%) | 0.402 |
| Dry cough, n/% | 3(50%) | 21(87.50%) | 17(73.91%) | 0.127 |
| Fatigue, n/% | 2(33.33%) | 9(37.50%) | 2(8.69%) | 0.062 |
| Chill, n/% | 1(16.67%) | 6(25.00%) | 3(13.04%) | 0.572 |
| Dyspnea, n/% | 1(16.67%) | 7(29.16%) | 3(13.04%) | 0.382 |
| Myalgia, n/% | 1(16.67%) | 3(12.5%) | 4(17.39%) | 0.890 |
| Expectoration, n/% | 3(50%) | 11(45.83%) | 11(47.83%) | 0.980 |
| Diarrhea, n/% | 2(33.33%) | 1(4.17%) | 5(21.74%) | 0.101 |
| Angina | 1(16.67%) | 3(12.5%) | 4(17.39%) | 0.890 |
| **Comorbidities** |  |  |  |  |
| Hypertension, n/% | 4(66.67%) | 2(8.33%)* | 2(8.69%)* | 0.001 |
| Diabetes, n/% | 2(33.33%) | 4(16.67%) | 2(8.69%) | 0.311 |
| Cardiovascular diseases, n/% | 4(66.67%) | 1(4.17%)* | 1(4.35%)* | <0.001 |
| COPD, n/% | 2(33.33%) | 0* | 1(4.35%)* | 0.025 |
| With more than two comorbidities, n/% | 5(83.33%) | 0* | 0* | <0.001 |
| CT characteristics of NCP |  |  |  |  |
| Bilateral involvement on CT or DR, n/% | 6(100%) | 21(87.5%) | 15(65.22%) | 0.070 |

Note: Data are presented as median (IQR) or n/%. \* mean $p < 0.05$ compared with the AMI group; # $p < 0.05$ compared with the group with cardiac marker abnormalities.
NCP, novel coronavirus pneumonia; T, temperature; HR, heart rate; BP, blood pressure; COPD, chronic obstructive pulmonary disease; IQR, interquartile range; AMI, acute myocardial injury; CT, computed tomography; DR, digital radiography.

High occurrence of hypertension was detected in the AMI patients (total patients with hypertension: n=8, 15.09%; AMI group: n=4, 66.67%; cardiac marker abnormalities group: n=2, 8.33%; without cardiac marker abnormalities group: n=2, 8.69%; p=0.001). Cardiovascular diseases were more frequently found in the AMI patients, all of whom had coronary artery disease, with one having prior coronary artery bypass grafting (total patients with cardiovascular disease: n=6, 11.32%; AMI group: n=4, 66.67%), compared with the groups with and without cardiac marker abnormalities (p=0.001). Additionally, diabetes (n=8, 15.09%) and COPD (n=3, 5.6%) were found in all the patients, with a higher tendency in the AMI group (diabetes: n=2 and COPD: n=2) than in the other two groups **(Figure 1)**. Notably, only the AMI patients had more than two comorbidities (n=5).

**Figure 1.**
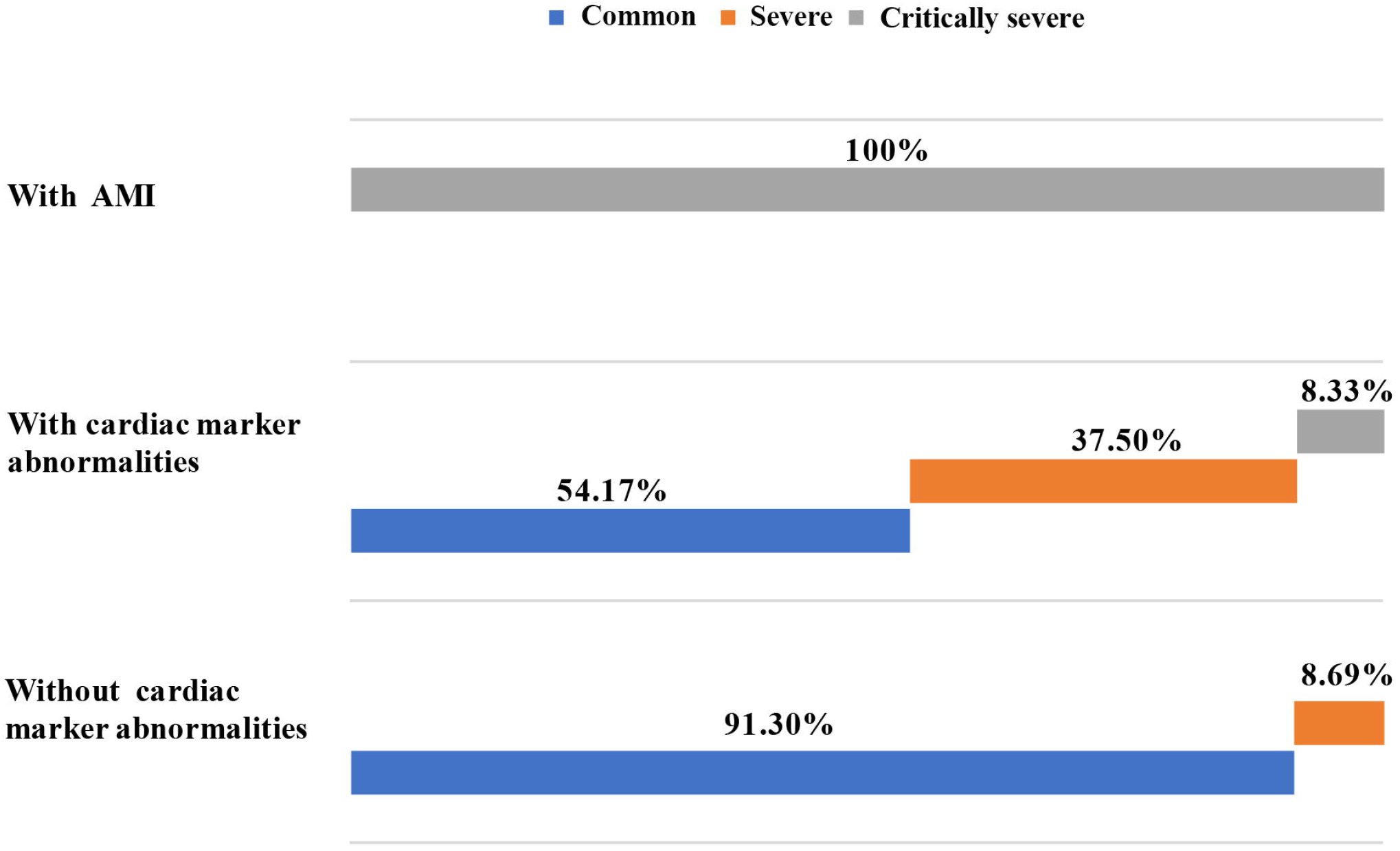
Comorbidities in the COVID-19 patients. In the AMI patients, hypertension and cardiovascular diseases were the frequently occurring comorbidities. AMI, acute myocardial injury; COPD, chronic obstructive pulmonary diseases; COVID-19, Coronavirus Disease 2019.

Regarding CT or DR findings on admission, all the AMI patients and 21 (87.5%) non-definite AMI patients with cardiac marker abnormalities had bilateral multilobular and sub-segmental lung involvement; in the without cardiac marker abnormalities group, bilateral lobes were affected in only 15 (65.22%) patients.

### Laboratory tests

The laboratory test and blood gas analysis results are shown in **Table S1 (see in supplementary appendix)**. CRP levels of all the AMI patients were increased above the normal range (0–5 mg/l) on admission and were higher than those of the other two groups; however, CRP levels were also increased in some patients in the groups with (n=9, 39.13%; CRP: median, 14.70 mg/ml [IQR 9.51, 24.53]) and without (n=4, 16.67%; CRP: median, 5.36 mg/ml [IQR 1.27,9.16]) cardiac marker abnormalities. D-dimer levels on admission were increased in the AMI patients (median, 1.65 µg/ml [IQR 1.23, 4.48]) and patients with abnormal cardiac markers (median, 0.79 µg/ml [IQR 0.62, 0.93]) and were higher than those in patients with normal cardiac markers (median, 0.55 µg/ml [IQR 0.46, 0.72]; p<0.001). Initial white blood cell counts of all the patients were within the normal range (3.50–9.50 × 10^9^ /l). Initial lymphocyte counts of five (83.33%) AMI patients were decreased below the normal range (0.80–4.00 × 10^9^ /l); these counts were decreased in only nine (37.5%) and eight (34.78%) patients with and without cardiac marker abnormalities, respectively. Furthermore, red blood cell count, hemoglobin level, and platelet count of the AMI group were lower than those of the other two groups (p<0.05); four (66.67%) AMI patients had procalcitonin levels above the normal range (0–0.5 ng/ml).

### Cardiac marker, echo, and ECG abnormalities

Over half (n=30, 56.6%) of the COVID-19 patients exhibited elevated cardiac marker levels. Cardiac marker levels were higher in the AMI and non-definite AMI patients than in those without cardiac marker abnormities (p<0.05; **Figure S1 in supplementary appendix)**. In the AMI group, cardiac marker levels continuously increased in three patients who died, with the levels being extremely high on the day of death **(Figure S2 in supplementary appendix)**.

Meanwhile, there were some differences in echo and ECG findings (**Table 2**). Of all the COVID-19 patients, 45.28% (n=24) patients exhibited echo abnormalities. Echo abnormalities more frequently occurred in the AMI group and included left atrial enlargement (n=2), left ventricular (LV) enlargement (n=2), LV diastolic dysfunction (n=3), mitral valve regurgitation (n=2), triple valve regurgitation (n=1), and aortic valve regurgitation (n=1). Three AMI patients had LV wall thickening. It is noteworthy that non-definite AMI patients with cardiac marker abnormalities also had high frequency of echo abnormalities (n=15, 62.50%), with 11(45.83%) of them having more than two of the abovementioned echo abnormalities. The most frequent echo abnormality in the cardiac marker abnormalities group was LV diastolic dysfunction (n=14, 58.33%).

**Table 2.** Echocardiographic and electrocardiographic result during admission.

| AMI group (n=6) | Cardiac marker | Without cardiac marker | p |
| --- | --- | --- | --- |

|  |  | abnormalities group (n=24) | abnormalities group (n=23) |  |
| --- | --- | --- | --- | --- |
| **Echo abnormalities** | 6(100%) | 15(62.50%)* | 3(13.04%)*# | <0.001 |
| LV enlargement, n/% | 2(33.33%) | 1(4.17%)* | 0* | 0.006 |
| LA enlargement, n/% | 2(33.33%) | 4(16.67%) | 0*# | 0.038 |
| LV diastolic dysfunction, n/% | 3(50%) | 14(58.33%) | 3(13.04%)*# | 0.005 |
| Mitral valve regurgitation, n/% | 2(33.33%) | 7(29.16%) | 0*# | 0.015 |
| Triple valve regurgitation, n/% | 1(16.67%) | 2(8.33%) | 0* | 0.216 |
| Aortic valve regurgitation, n/% | 1(16.67%) | 3(12.5%) | 0* | 0.179 |
| LV wall thickening, n/% | 3(50%) | 4(16.67%) | 0*# | 0.004 |
| With two items, n/% | 6(100%) | 11(45.83%)* | 0*# | <0.001 |
| **ECG abnormalities** | 6(100%) | 5(20.83%)* | 0*# | <0.001 |
| ARB, n/% | 2(33.33%) | 0* | 0* | <0.001 |
| ST-T/Q curve abnormalities, n/% | 2(33.33%) | 5(20.83%) | 0*# | 0.033 |
| Arrhythmia, n/% | 3(50%) | 1(4.27%)* | 0* | <0.001 |
| With two items, n/% | 5(83.33%) | 1(4.27%)* | 0* | <0.001 |

Note: Data are presented as n (%). \* mean p<0.05 compared with the AMI group; # p<0.05 compared with the group with cardiac marker abnormalities.
LV, left ventricle; LA, left atrium; Echo, echocardiography; ECG, electrocardiogram; ARB, atrioventricular block; other abbreviations are the same as those in the footnote of Table 1.

As shown in Table 3, 11 (20.75%) COIVD-19 patients had ECG abnormalities, including all the AMI patients and five (20.83%) non-definite AMI patients with cardiac marker abnormalities. Five of the AMI patients had more than two kinds of ECG abnormalities including atrioventricular block (n=2), ST-T/Q curve abnormalities (n=2), and arrhythmia (n=3); furthermore, ST-T/Q curve abnormalities (n=5, 20.83%) and arrhythmia (n=1, 4.27%) were the main abnormalities in patients with cardiac marker abnormalities. None of the patients without cardiac marker abnormalities exhibited ECG abnormalities.

**Table 3.**
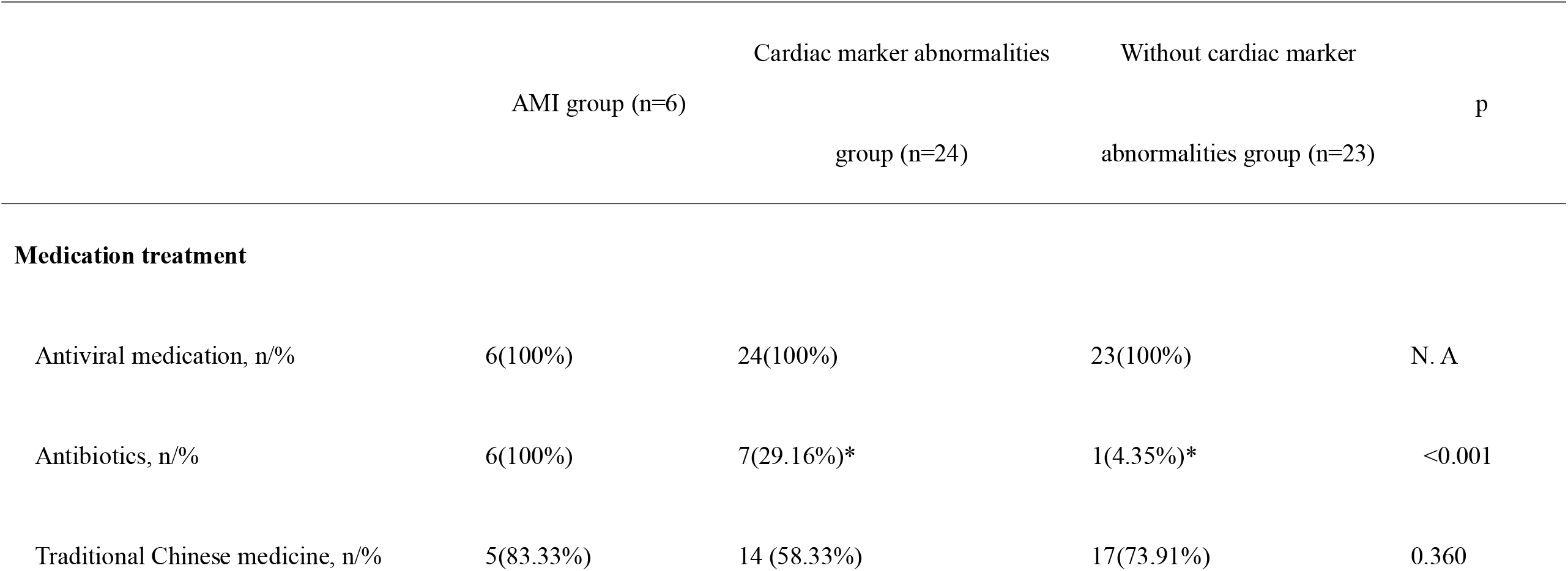
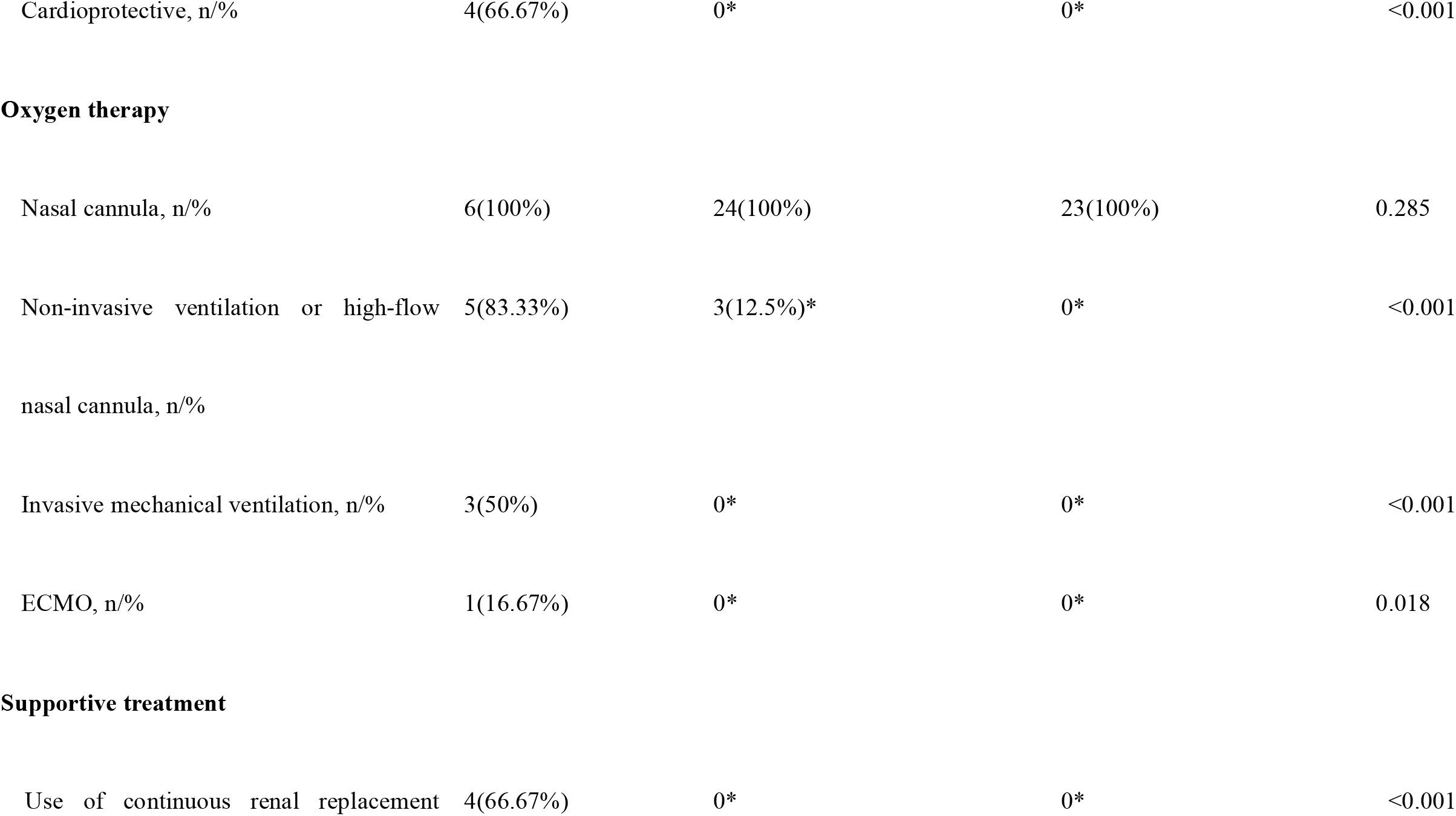
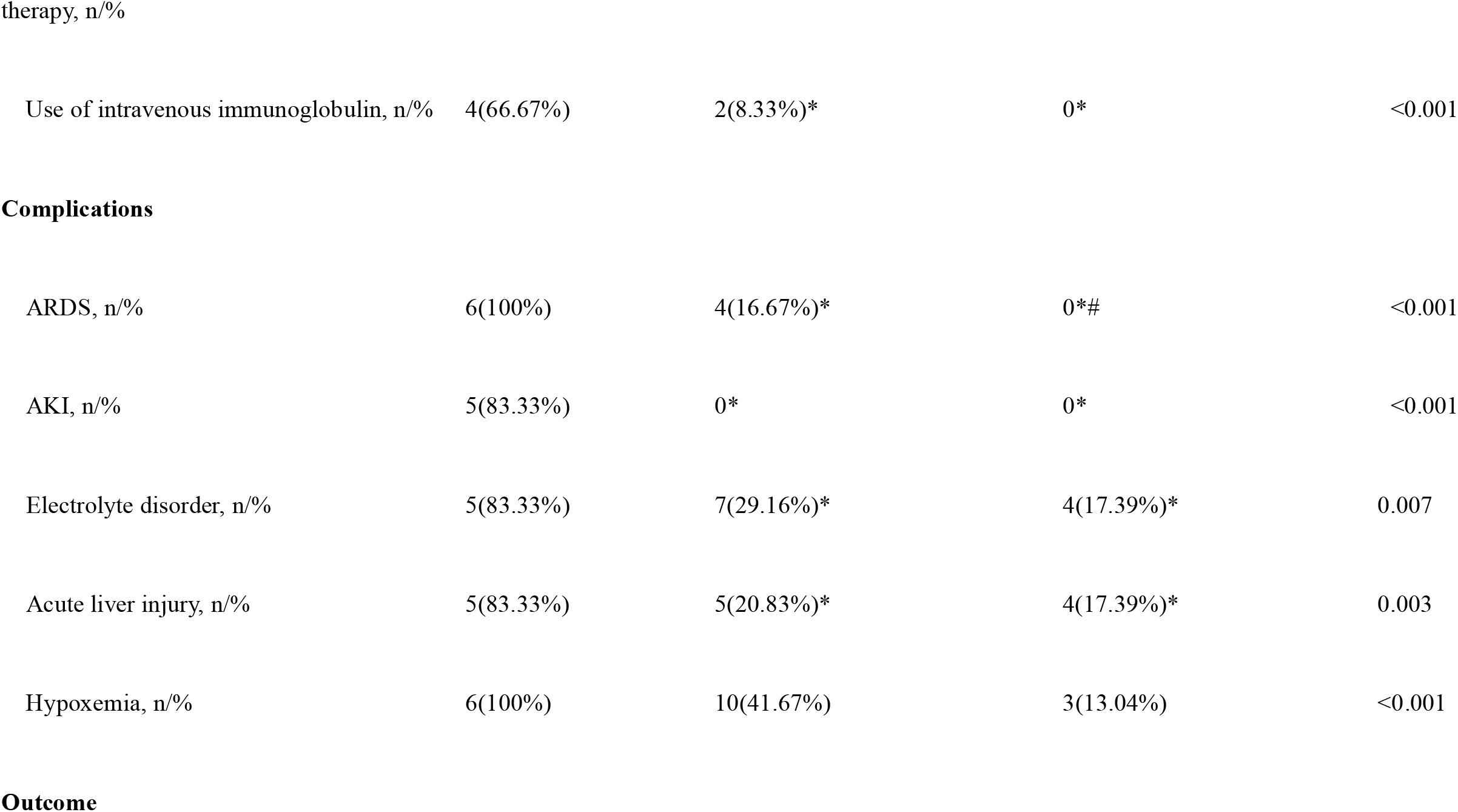
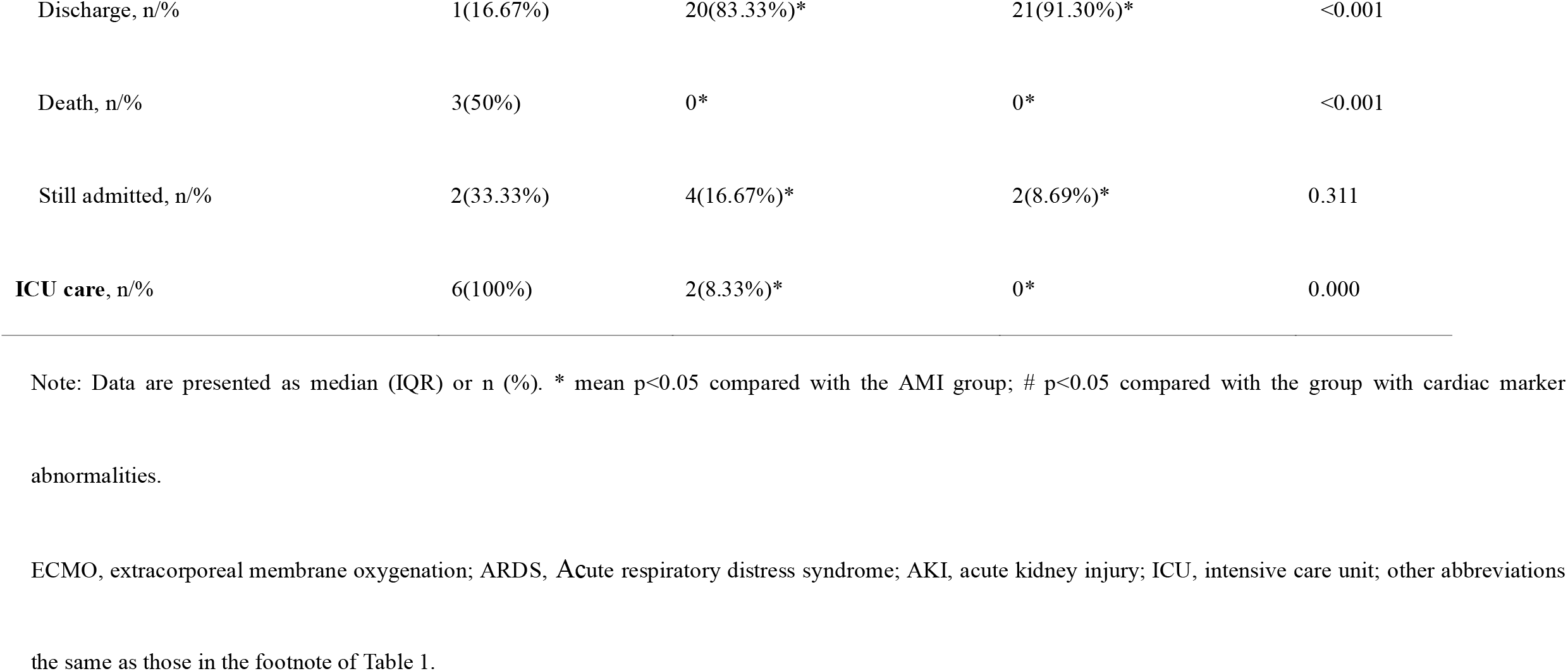
Main management strategies, comorbid conditions, and clinical outcomes.

### Main management strategies, comorbid conditions, and clinical outcome data

**Table 3** shows the main management strategies, comorbid conditions, and clinical outcomes of all the COVID-19 patients. In the AMI group, in addition to antiviral and antibiotic therapies, cardioprotective medications such as amiodarone and cedilla were used by four patients to improve the cardiac function. However, the non-definite AMI patients with and without cardiac markers abnormalities did not use cardioprotective medication. All the 53 patients were administered nasal cannula oxygen therapy during admission for respiratory support; non-invasive ventilation or high-flow nasal cannula oxygen therapy was performed for five AMI and three non-definite AMI patients with cardiac marker abnormalities. Notably, in the AMI group, invasive mechanical ventilation was used in three patients and ECMO in one patient due to extremely severe ARDS.

ARDS occurred in all the six AMI patients and four non-definite AMI patients with cardiac marker abnormalities. Hypoxemia was a common complication that affected 19 (35.85%) of the 53 study patients, with a significant difference in the number of affected patients between the AMI (n=6, 100%) and non-definite AMI with cardiac marker abnormalities (n=10, 41.67%) groups and the without cardiac marker abnormalities group (n=3, 13.04%; p<0.001). Moreover, a total of five AMI patients had AKI, and four of them used continuous renal replacement therapy.

Importantly, all the six AMI patients had critically severe NCP and needed care in intensive care unit (ICU). However, half of the patients with cardiac marker abnormalities (n=13, 54.17%) had the common NCP type, with only two (8.33%) of them having critically severe NCP needing ICU care. Meanwhile, the majority of patients (n=21, 91.30%) in the without cardiac marker abnormalities group had the common NCP type (**Figure 2**). Regarding clinical outcomes, in the AMI group, three patients died, two were still in admission, and one was discharged from hospital. Conversely, a majority of non-definite AMI patients with (n=20, 83.33%) and without (n=21, 91.30%) cardiac marker abnormalities were discharged, with no case of death.

**Figure 2.**
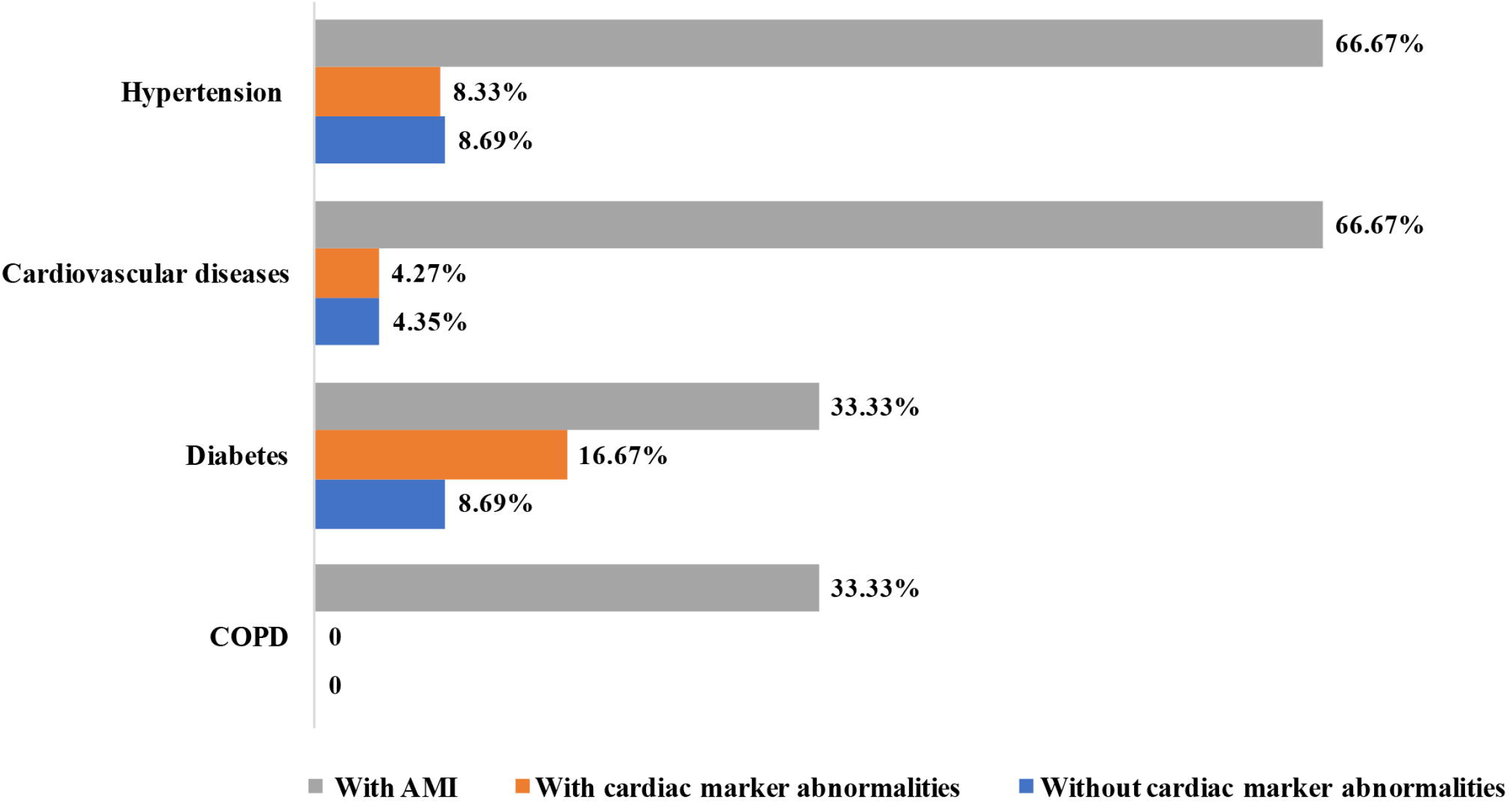
Heterogeneity of NCP types in the COVID-19 patients. NCP, novel coronavirus pneumonia. Abbreviations are the same as those for

### Risk factors for AMI

To determine the risk factors for AMI in the COVID-19 patients, a bivariate analysis of AMI occurrence with clinical data and laboratory findings was performed, showing that age (r=0.456, p=0.001), CRP levels (r=0.483, p<0.001), NCP severity (r=0.540, p<0.001), and presence of commodities (r=0.520, p<0.001) were associated with AMI occurrence (all p<0.05). Further analysis (**Table S2 in supplementary appendix**) demonstrated that increased CRP levels (OR, 1.186; 95CI% [1.047, 1.344]), representing the inflammatory condition of the body, may be a risk factor for AMI in COVID-19 patients. COVID-19 patients with more severe NCP type (OR, 2.896; 95CI% [1.266, 6.621]) may have a higher risk of AMI than other patients. It is worth mentioning that the presence of comorbidities (OR, 4.336; 95CI% [1.11, 38.55]) may be the most critical risk factor for AMI in COVID-19 patients.

## Discussion

This is the first study to show that cardiac abnormalities including tachycardia (28%), elevated myocardial enzyme levels (56.6%), cardiac dysfunction (37.7%), and even AMI (11.3%) are common in COVID-19 patients. Moreover, AMI was detected in 6 of the 53 hospitalized COVID-19 patients. All the six patients had severe or critically severe NCP and required ICU care. Of the six AMI patients, three died, two remained hospitalized, and one was discharged. Notably, CRP levels, NCP severity, and underlying comorbidities were the major risk factors for AMI in the COVID-19 patients. Interestingly, there were 24 (45%) COVID-19 patients with elevation in the levels of one or more myocardial enzymes and markers of myocardial damage whose features did not meet the strict definition of myocardial injury, with most of them having the common NCP type and experiencing quick recovery; however, 58.33% of these patients exhibited LV dysfunction on echo, and long-term outcomes should be observed via follow-up in a future study.

Our study results suggest that the rates of cardiac complications were high in the hospitalized COVID-19 patients, especially in patients with confirmed AMI from among those who required ICU care. As shown in a recent report on 138 hospitalized COVID-19 patients,^2^ 16.7% of the patients developed arrhythmia and 7.2% developed acute cardiac injury. In another series of 41 cases of hospitalized COVID-19 patients, it has been reported that 5 (12%) patients were diagnosed with acute cardiac injury.^2-5^ Similar to the cardiovascular complications of COVID-19, those of SARS include hypotension, tachycardia, arrhythmias, systolic and diastolic dysfunctions, and sudden death, with tachycardia particularly being the commonest condition persistent in nearly 40% of patients during follow-up.^14^ Furthermore, MERS-induced myocarditis and myocardial damage has been reported.^15^ However, case series reporting elevated levels of myocardial enzymes and myocardial injury markers remain limited for SARS and MERS. Notably, although only six patients had clinically confirmed AMI based on strict inclusion criteria in our study, most patients had elevated levels of one or more cardiac injury biomarkers and LV diastolic dysfunction. Although long-term outcomes of COVID-19 patients with AMI need to be further validated, significantly more clinical attention should be paid to avoid underdiagnosis because of classic symptoms and because of the possible overshadowing of cardiac complications in the context of coronavirus, as cautioned by the ACC Clinical Bulletin.^16^

To date, the underlying mechanisms of AMI and potential impacts in COVID-19 patients remain unknown. Clinically, the increased serum levels of proinflammatory cytokines, such as interleukin (IL)-1β, IL-6, IL-12, interferon-gamma, interferon-inducible protein 10, and monocyte chemoattractant protein-1, suggest that severe acute respiratory syndrome coronavirus 2 (SARS-CoV-2) infection is associated with cellular immune deficiency and coagulation activation.^17^ Thus, the direct effects of the virus, cytokine storm induced by viral invasion, and sustained inflammatory response are presumed to be the main pathophysiological mechanisms underlying SARS-CoV-2 infection. Recently, a case report showing pathological findings from a severely infected COVID-19 patient revealed no obvious histological changes in heart tissue; however, the conclusion that SARS-CoV-2 infection might not directly impair the heart is debatable because this finding was observed in only one patient without AMI-related clinical evidences.^18^ Because the three patients in our study who died all had extremely elevated cardiac marker levels, acute heart failure may play a fatal role in multiple organ dysfunction and accelerate death. However, further evidence is urgently needed to assess the mechanism of cardiac damage in large-sample autopsy or biopsy studies. Previous studies have indicated that SARS-CoV can mediate MI and damage associated with the downregulation of the myocardial angiotensin-converting enzyme 2 system; this may be responsible for the myocardial dysfunction and adverse cardiac outcomes in patients with SARS, which has also been detected in COVID-19 patients.^19-22^ Until now, the pathophysiology of SARS-CoV or MERS-CoV has not been completely understood. Although full-genome sequencing and phylogenic analysis has shown that SARS-CoV-2 is similar to SARS-CoV or MERS-CoV, the pathophysiological mechanism of cardiac infection or damage caused by SARS-CoV-2 needs to be further validated in future studies.^23^

In our study, AMI usually occurred in COVID-19 patients with old age, underlying comorbidities, severe or critically severe NCP, and ARDS. Among 44,672 patients with confirmed COVID-19, as reported in the China CDC Weekly on Feb 11, 2020, approximately 31.2% patients were aged >60 years. The overall case fatality rate was 2.3% (1,023 deaths); more importantly, the majority (81%) of deaths occurred in patients aged ≥60 years or in those with underlying medical conditions.^24^ Similarly, in our study, all the six AMI patients were aged >60 years and had one or more underlying conditions, including diabetes, hypertension, COPD, and cardiovascular diseases. As described in a retrospective study of 1,099 laboratory-confirmed cases, approximately 25.2% of the patients had at least one underlying disorder, such as hypertension and COPD, and an underlying disorder potentially acts as an important risk factor for poor outcomes.^25^ We found that the severity of NCP was closely correlated with the AMI occurrence. Meanwhile, ARDS was present in all the AMI patients. ARDS, as a severe complication, occurred in approximately 19%–29% NCP patients as per previous reports.^2-5^ Thus, we must understand the mechanisms of heart–lung interactions in ARDS patients, particularly the right ventricle function impairments caused by respiratory dysfunction.^26^ Logistic regression analysis indicated that NCP severity was a risk factor for AMI in COVID-19 patients. Indeed, the outcomes of these patients need to be further determined in long-term follow-up studies.

Our study has some limitations. First, this is a modest-size case series of hospitalized COVID-19 patients; more standardized data from a larger cohort would beneficial for further determining the clinical characteristics. Second, our data showed the clinical characteristics of AMI induced by COVID-19, including the clinical presentations, electrophysiological abnormality, myocardial enzyme and myocardial injury marker levels, and cardiac dysfunction and enlargement. However, the tissue characteristics in myocardial damage (i.e., edema, fibrosis, and microcirculation disorder) should be further demonstrated via cardiac magnetic resonance imaging examination as far as possible under the premise of controlling transmission. Finally, most of the COVID-19 patients with AMI had severe or critically severe NCP and high fatality rate, but long-term follow-up should be performed to determine adverse cardiac events in the patients who could not be diagnosed with MI but had elevated levels of one or more myocardial enzymes and myocardial injury markers.

In summary, we found that cardiovascular complications are common in COVID-19 patients and include tachycardia, elevated myocardial enzyme levels, cardiac dysfunction, and even AMI. More importantly, CRP level elevation, NCP severity, and underlying cardiovascular diseases are the major risk factors for AMI in these patients.

## Data Availability

All data referred to in the manuscript are available.

## Acknowledgments

We thank all our colleagues who helped us during the current study. We are also grateful to the many front-line medical sta□ for their dedication in the face of this outbreak, despite the potential threat to their own lives and the lives of their families.

## Funding

This work was supported by 2020 Novel coronavirus pneumonia prevention and control technology project of Chengdu (NO. 2020-YF05-00007-SN); Clinical research finding of Chinese society of cardiovascular disease(CSC) of 2019 (NO. HFCSC2019B01); the National Natural Science Foundation of China (NO. 81771887, 81771897, 81971586,81901712); the Program for Young Scholars and Innovative Research Team in Sichuan Province (NO. 2017TD0005) of China; and 1·3·5 project for disciplines of excellence, West China Hospital, Sichuan University (NO.ZYGD18013).

## Conflict of interest disclosures

none

## Notes

Conflict of Interest: none declared

### Competing Interest Statement

The authors have declared no competing interest.

### Clinical Trial

This article is a retrospective research.

## Reference

1. Coronavirus disease 2019 (COVID-19) situation report-43. WHO. https://www.who.int/docs/default-source/coronaviruse/situation-reports/20200303-sitrep-43-covid-19.pdf?sfvrsn=2c21c09c_2

2. Wang D, Hu B, Hu C, Zhu F, Liu X, Zhang J, Wang B, Xiang H, Cheng Z, Xiong Y, Zhao Y, Li Y, Wang X, Peng Z. Clinical Characteristics of 138 Hospitalized Patients With 2019 Novel Coronavirus-Infected Pneumonia in Wuhan, China. JAMA. 2020 Feb 7. doi: 10.1001/jama.2020.1585.

3. Huang C, Wang Y, Li X, Ren L, Zhao J, Hu Y, Zhang L, Fan G, Xu J, Gu X, Cheng Z, Yu T, Xia J, Wei Y, Wu W, Xie X, Yin W, Li H, Liu M, Xiao Y, Gao H, Guo L, Xie J, Wang G, Jiang R, Gao Z, Jin Q, Wang J, Cao B. Clinical features of patients infected with 2019 novel coronavirus in Wuhan, China. Lancet. 2020;395(10223):497–506.

4. Na Zhu, Dingyu Zhang, Wenling Wang, Xingwang Li, Bo Yang, Jingdong Song, Xiang Zhao, Baoying Huang, Weifeng Shi, Roujian Lu, Peihua Niu, Faxian Zhan, Xuejun Ma, Dayan Wang, Wenbo Xu, Guizhen Wu, George F. Gao, D. Phil, Wenjie Tan. for the China Novel Coronavirus Investigating and Research Team. A novel coronavirus from patients with pneumonia in China, 2019. https://www.nejm.org/doi/full/10.1056/NEJMoa2001017?query=recirc_mostViewed_railB_article

5. Chen N, Zhou M, Dong X, Qu J, Gong F, Han Y, Qiu Y, Wang J, Liu Y, Wei Y, Xia J, Yu T, Zhang X, Zhang L. Epidemiological and clinical characteristics of 99 cases of 2019 novel coronavirus pneumonia in Wuhan, China: a descriptive study. Lancet. 2020;395(10223):507–513. doi: 10.1016/S0140-6736(20)30211-7.

6. Newby LK, Jesse RL, Babb JD, Christenson RH, De Fer TM, Diamond GA, Fesmire FM, Geraci SA, Gersh BJ, Larsen GC, Kaul S, McKay CR, Philippides GJ, Weintraub WS. ACCF 2012 expert consensus document on practical clinical considerations in the interpretation of troponin elevations: a report of the American College of Cardiology Foundation task force on Clinical Expert Consensus Documents. J Am Coll Cardiol. 2012;60(23):2427–63.

7. Li SS, Cheng CW, Fu CL, Chan YH, Lee MP, Chan JW, Yiu SF. Left ventricular performance in patients with severe acute respiratory syndrome: a 30-day echocardiographic follow-up study. Circulation. 2003;108(15):1798–803.

8. Alexander LK, Keene BW, Small JD, Yount B Jr, Baric RS. Electrocardiographic changes following rabbit coronavirus-induced myocarditis and dilated cardiomyopathy. Adv Exp Med Biol. 1993;342:365–70.

9. Yu CM, Wong RS, Wu EB, Kong SL, Wong J, Yip GW, Soo YO, Chiu ML, Chan YS, Hui D, Lee N, Wu A, Leung CB, Sung JJ. Cardiovascular complications of severe acute respiratory syndrome. Postgrad Med J. 2006 Feb;82(964):140–4.

10. Madjid M, et al. ACC Clinical Bulletin: Cardiac Implications of Novel Wuhan Coronavirus (2019-nCoV). Accessed Feb. 13, 2020.

11. National Health Commission of the People’s Republic of China. Diagnosis and treatment protocols of pneumonia caused by a novel coronavirus (Revised version 5). Published on February 8, 2020. http://www.nhc.gov.cn/xcs/zhengcwj/202002/d4b895337e19445f8d728fcaf1e3e13a.shtml.

12. ARDS Definition Task Force, Ranieri VM, Rubenfeld GD, Thompson BT, Ferguson ND, Caldwell E, Fan E, Camporota L, Slutsky AS. Acute respiratory distress syndrome: the Berlin Definition. JAMA. 2012;307(23):2526–33.

13. Kidney Disease: Improving Global Outcomes (KDIGO) Acute Kidney Injury Work Group. KDIGO Clinical Practice Guideline for Acute Kidney Injury. Kidney Int Suppl. 2012;2:1.

14. Ding Y, He L, Zhang Q, Huang Z, Che X, Hou J, Wang H, Shen H, Qiu L, Li Z, Geng J, Cai J, Han H, Li X, Kang W, Weng D, Liang P, Jiang S. Organ distribution of severe acute respiratory syndrome (SARS) associated coronavirus (SARS-CoV) in SARS patients: implications for pathogenesis and virus transmission pathways. J Pathol. 2004;203(2):622–30.

15. Alhogbani T. Acute myocarditis associated with novel Middle east respiratory syndrome coronavirus. Ann Saudi Med. 2016;36(1):78–80.

16. Madjid M, et al. ACC Clinical Bulletin: Cardiac Implications of Novel Wuhan Coronavirus (2019-nCoV). Accessed Feb. 13, 2020.

17. Chaolin Huang, Yeming Wang, Xingwang Li, Lili Ren, Jianping Zhao, Yi Hu, Li Zhang, Guohui Fan, Jiuyang Xu, Xiaoying Gu, Zhenshun Cheng, Ting Yu, Jiaan Xia, Yuan Wei, Wenjuan Wu, Xuelei Xie, Wen Yin, Hui Li, Min Liu, Yan Xiao, Hong Gao, Li Guo, Jungang Xie, Guangfa Wang, Rongmeng Jiang, Zhancheng Gao, Qi Jin, Jianwei Wang, Bin Cao. Clinical features of patients infected with 2019 novel coronavirus in Wuhan, China. Lancet. January 24, 2020 https://doi.org/10.1016/S0140-6736(20)30183-5.

18. Zhe Xu, Lei Shi, Yijin Wang, Jiyuan Zhang, Lei Huang, Chao Zhang, Shuhong Liu, Peng Zhao, Hongxia Liu, Li Zhu, Yanhong Tai, Changqing Bai, Tingting Gao, Jinwen Song, Peng Xia, Jinghui Dong, Jingmin Zhao, Fu-Sheng Wang. Pathological findings of COVID-19 associated with acute respiratory distress syndrome. The Lancet Respiratory Medicine. Published: February 18, 2020 DOI:https://doi.org/10.1016/S2213-2600(20)30076-X.

19. Kuba K, Imai Y, Rao S, Gao H, Guo F, Guan B, Huan Y, Yang P, Zhang Y, Deng W, Bao L, Zhang B, Liu G, Wang Z, Chappell M, Liu Y, Zheng D, Leibbrandt A, Wada T, Slutsky AS, Liu D, Qin C, Jiang C, Penninger JM. A crucial role of angiotensin converting enzyme 2 (ACE2) in SARS coronavirus-induced lung injury. Nat Med. 2005;11(8):875–9.

20. Oudit GY, Kassiri Z, Jiang C, Liu PP, Poutanen SM, Penninger JM, Butany J. SARS-coronavirus modulation of myocardial ACE2 expression and inflammation in patients with SARS. Eur J Clin Invest. 2009;39(7):618–25.

21. Hamming I, Timens W, Bulthuis ML, Lely AT, Navis G, van Goor H. Tissue distribution of ACE2 protein, the functional receptor for SARS coronavirus. A first step in understanding SARS pathogenesis. J Pathol. 2004;203(2):631–7.

22. Wu, C, Zheng, M. Single-Cell RNA Expression Profiling Shows that ACE2, the Putative Receptor of Wuhan 2019-nCoV, Has Significant Expression in the Nasal, Mouth, Lung and Colon Tissues, and Tends to be Co-Expressed with HLA-DRB1 in the Four Tissues. Preprints 2020, 2020020247.

23. de Wit E, van Doremalen N, Falzarano D, Munster VJ. SARS and MERS: recent insights into emerging coronaviruses. Nat Rev Microbiol. 2016;14 (8):523–534. doi:10.1038/nrmicro.2016.81.

24. The Novel Coronavirus Pneumonia Emergency Response Epidemiology Team. The Epidemiological Characteristics of an Outbreak of 2019 Novel Coronavirus Diseases (COVID-19) — China, 2020. China CDC Weekly, 2020, 2(8): 113–122. http://weekly.chinacdc.cn/en/article/id/e53946e2-c6c4-41e9-9a9b-fea8db1a8f51.

25. Wei-jie Guan, Zheng-yi Ni,Yu Hu, Wen-hua Liang, Chun-quan Ou, Jian-xing He, Lei Liu, Hong Shan, Chun-liang Lei, David S.C. Hui, Bin Du, Lan-juan Li, Guang Zeng, Kwok-Yung Yuen, Ru-chong Chen,Chun-li Tang, Tao Wang, Ping-yan Chen, Jie Xiang, Shi-yue Li, Jin-lin Wang, Zi-jing Liang, Yi-xiang Peng, Li Wei, Yong Liu, Ya-hua Hu, Peng Peng, Jian-ming Wang, Ji-yang Liu, Zhong Chen, Gang Li, Zhi-jian Zheng, Shao-qin Qiu, Jie Luo, Chang-jiang Ye, Shao-yong Zhu, and Nan-shan Zhong. for the China Medical Treatment Expert Group for Covid-19. Clinical Characteristics of Coronavirus Disease 2019 in China. February 28, 2020.DOI: 10.1056/NEJMoa2002032.

26. Yu CM, Wong RS, Wu EB, Kong SL, Wong J, Yip GW, Soo YO, Chiu ML, Chan YS, Hui D, Lee N, Wu A, Leung CB, Sung JJ. Cardiovascular complications of severe acute respiratory syndrome. Postgrad Med J. 2006;82(964):14.

